# COVID-19-related Smell and Taste Impairment with Widespread Diffusion of SARS-CoV-2 Omicron Variant

**DOI:** 10.1101/2022.02.17.22271116

**Authors:** Paolo Boscolo-Rizzo, Giancarlo Tirelli, Pierluigi Meloni, Claire Hopkins, Giordano Madeddu, Andrea De Vito, Nicoletta Gardenal, Romina Valentinotti, Margherita Tofanelli, Daniele Borsetto, Jerome R. Lechien, Jerry Polesel, Giacomo De Riu, Luigi Angelo Vaira

## Abstract

**Background:** The aim of this study was to estimate the prevalence of self-reported chemosensory dysfunction in a study cohort of subjects who developed a mild-to-moderate COVID-19 in the period from January 17, 2022 to February 4, 2022 (Omicron proxy period) and compared that with a historical series of patients tested positive for SARS-CoV-2 infection between March and April, 2020 (comparator period).

**Methods:** Prospective study based on the sinonasal outcome tool 22 (SNOT-22), item “sense of smell or taste” and additional outcomes.

**Results:** Patients’ characteristics and clinical presentations of COVID-19 were evaluated and compared in 779 patients, 338 of the study cohort and 441 of the historical series. The prevalence of self-reported chemosensory dysfunction during the proxy Omicron period (32.5%; 95% CI, 27.6-37.8) was significantly lower from that during the comparator period (66.9%; 95% CI, 62.3-71.3) (*p*<.001). 24.6% (95% CI, 20.1-29.5) of patients reported an altered sense of smell during the proxy Omicron period compared to 62.6% (95% CI, 57.9-67.1) during the comparator period (*p*<.001). Similarly, the prevalence of an altered sense of taste dropped from 57.4% (95% CI, 52.6-62.0) during the comparator period to 26.9% (95% CI, 22.3-32.0) during the proxy Omicron period (*p*<.001). The severity of chemosensory dysfunction was lower in proxy Omicron period compared to comparator period (*p*<.001).

**Conclusions:** The prevalence and the severity of COVID-19 associated smell and taste dysfunction has dropped significantly with the advent of the Omicron variant.

## Introduction

In December 2021, WHO defined five severe acute respiratory coronavirus 2 (SARS-CoV-2) variants of concern (VOC), Alpha (B.1.1.7), Beta (B.1.351), Gamma (P.1), Delta (B.1.617.2 and AY lineages) and Omicron (originally B.1.1.529, then reclassified into BA lineages) ^1^. The Omicron variant, first detected in South Africa on October 24, 2021, represents the most recently recognized VOC ^2^. Compared to Delta, which was the most prevalent variant worldwide in December 2021, Omicron spread more rapidly becoming the dominant variant in January 2022 ^3^.

Omicron seems to cause a less severe disease with determinants of severity being multifactorial and including a lower replication competence in the lung parenchyma compared to bronchus ^4^. Consistently, the spectrum of symptoms is expected to differ from that observed in the coronavirus disease 2019 (COVID-19) driven by other SARS-CoV-2 strains. However, to the best of our knowledge, only one report has been published so far regarding the prevalence of different symptoms in infections driven by this VOC ^5^.

Smell and taste dysfunction were consistently reported among the most common symptoms of COVID-19 with about 65-70% of patients with mild-to-moderate disease experiencing a chemosensory impairment during the acute phase of the COVID-19 ^6–8^. Recently, in a series of 81 subjects tested positive for the SARS-CoV-2 Omicron variant, the impairment of the sense of smell and taste was self-reported by 12% and 23% of patients, respectively ^5^. Since January 17, 2022, Omicron variant was by far the most predominant variant in Italy with an overall prevalence of 95.8% ^9^. Particularly, in Friuli Venezia-Giulia and Sardinia, the prevalence of SARS-CoV-2 infection driven by Omicron variant was 97.0% and 96.2%, respectively ^9^.

The aim of this study was to determinate the prevalence of self-reported chemosensory dysfunction in a series of Italian subjects who developed a mild-to-moderate COVID-19 after January 17, 2022 and to compare it with that of a cohort of patients who tested positive for SARS-CoV-2 infection and were evaluated during the first wave of the pandemic in Italy.

## Materials and Methods

The study was approved by the Ethics Committees of the Friuli Venezia Giulia Region (CEUR-OS156) and University Hospital of Cagliari (PG 2021/7118). Informed consent was obtained for telephone interviews.

### Subjects

This is a prospective study on mild-to-moderate symptomatic adult patients resident in Friuli Venezia Giulia and Sardinia, who tested positive for SARS-CoV-2 RNA by polymerase chain reaction (PCR) on nasopharyngeal swabs performed according to World Health Organization recommendation between January 17 and February 4, 2022 ^10^. Consecutive contacts of subjects with a confirmed diagnosis of SARS-CoV-2 infection were identified by the hospitals involved. Patients were considered mildly-to-moderately symptomatic if they had less severe clinical symptoms with no evidence of pneumonia, not requiring hospitalization, and therefore considered suitable for being treated at home. Participants had to be interviewed within one month of the first positive swab. To be included in the study, subjects had to be recovered from the infection with a negative PCR confirmation on the nasopharyngeal swab or have had remission of symptoms for at least 7 days. The exclusion criteria were: contact information not available, uncooperative patients, assisted ventilation, psychiatric or neurological disorders, previous surgery or radiotherapy in the oral and nasal cavities, pre-existing self-reported smell and taste dysfunction, history of head trauma, allergic rhinitis, and chronic rhinosinusitis. The subjects were contacted by telephone by the researchers and interviewed.

### Questionnaires

Telephone interview were conducted between January, 28 and February 14, 2022. Demographic and clinical data were collected through standardized questions administered during the interview including gender, age, self-reported height and weight, smoking habit, and the following co-morbidities: immunosuppression, diabetes, cardiovascular diseases, active cancer, chronic respiratory disease, kidney disease, liver disease. Obesity was defined as having a body mass index (BMI) of 30 or more. Symptoms were assessed through standardized questions and structured questionnaires, including the Acute Respiratory Tract Infection Questionnaire (ARTIQ; with symptoms scored as none, 0; a little, 1; a lot, 2) and the Sino-Nasal Outcome test 22 (SNOT-22), item “sense of smell or taste” as previously reported ^6^. The SNOT-22 ranks symptom severity as none (0), very mild (1), mild or slight (2), moderate (3), severe (4), or as bad as it can be (5). Patients with SNOT-22≥1 were also asked, based on a binary outcome of yes and no, whether the chemosensory dysfunction involved the sense of smell, taste, or both. Then, patients were asked whether their gustatory alteration involved the perception of basic taste (“Do you have an impairment in the perception of fine taste, e.g., during eating and drinking?) or flavour (“Do you have an impairment in the perception your basic taste: sweet, sour, salty, bitter?). The dates of the first positive and negative swabs were obtained. In addition, patients were asked if they had already been infected with SARS-CoV-2 since the beginning of the pandemic and if they had been vaccinated and with how many doses. Individuals were considered fully vaccinated if they had received the required dose(s) of a SARS-CoV-2 vaccine and were at least 14 days after completion.

### Statistical analysis

We compared demographic and clinical data, with special emphasis to chemosensory dysfunction, of patients who developed COVID-19 in the period from January 17, 2022 to February 4, 2022 in Italy (Omicron proxy period), with an historical cohort of patients who complete the same outcomes prospectively, resident in the same Italian regions, who developed COVID-19 between March and April, 2020 ^11–14^, when G614 variant^15^ was dominant (comparator period). Symptom prevalence was expressed as percentage of total patients, and 95% confidence interval (CI) were calculated using Clopper-Pearson method. Differences in prevalence were evaluated through Fisher’s exact test and odds ratios (ORs) for variables associated for chemosensory dysfunction were calculated according to multivariable unconditional logistic regression model adjusted for age and gender. Analyses were performed using R 3.6. and statistical significance was claimed for p<0.05 (two-tailed).

## Results

The study included 779 patients, 338 from the study cohort (proxy Omicron period) and 441 from the control cohort (comparator period).

### Characteristics of the study cohort (proxy Omicron period)

Of 482 potential eligible patients, 144 did not respond or decline to take part in the survey leaving a total of 338 (70.1%; median [IQR] age, 46 [34-59] years; 183 [54%] women) who participated in the study. Patients’ characteristics are reported in **Table 1**. Associated co-morbidities were reported by 116 subjects (34.3%) with the most common being cardiovascular diseases reported by 56 patients (16.6%). A total of 279 patients (82.5%) reported that they had been fully vaccinated for SARS-CoV-2. 18 patients (5.3%) reported having already contracted a SARS-CoV-2 infection during the previous two years. Most frequent symptoms were blocked nose (68.3%), fever (58.9%), and dry cough (56.8%) (**Table 2**). Alterations of sense of smell or taste were reported by 110 patients (32.5%, 95% CI 27.6-37.8), with 61 patients reporting a SNOT-22>2 (18.0; 95% CI: 14.1.-22.6). Eighteen patients (5.3%) reported a score of 5 (**Table 3**). When asked about basic taste and flavour perception, 72 (21.3%) and 87 (25.7%) patients, respectively, self-reported an impairment with 68 (20.1%) subjects reporting both (data not shown).

**Table 1.** Baseline characteristics of patients with mild-to-moderate COVID-19 during the proxy Omicron period versus comparator period.

|  | Proxy Omicron period<br>(n=338) |  | Comparator period<br>(n=441) |  |  |
| --- | --- | --- | --- | --- | --- |
|  | n | Prevalence<br>% (95% CI)<sup>a</sup> | n | Prevalence<br>% (95% CI)<sup>a</sup> | *p-value* |
| Age, years (median, range) | 46 (34-59) |  | 50 (39-58) |  | 0.160 |
| Sex |  |  |  |  |  |
| Male | 155 | 45.9 (40.5-51.3) | 199 | 45.1 (40.4-51.4) | .885 |
| Female | 183 | 54.1 (48.7-559.5) | 242 | 54.9 (50.1-59.6) |  |
| Smoking status |  |  |  |  |  |
| Never | 200 | 59.2 (53.7-64.5) | 277 | 62.8 (58.1-67.3) | .335 |
| Ever | 138 | 40.8 (35.5-46.3) | 164 | 37.2 (32.7-41.9) |  |
| Current alcohol drinking |  |  |  |  |  |
| No | 245 | 72.4 (67.4-77.2) | 238 | 54.0 (49.2-58.7) | <.001 |
| Yes | 93 | 27.5 (22.8-32.6) | 203 | 46.0 (41.3-50.8) |  |
| Comorbidity |  |  |  |  |  |
| None | 222 | 65.7 (60.4-70.7) | 297 | 67.3 (62.8-71.7) | .008 |
| 1 | 68 | 20.1 (16.0-25.0) | 110 | 24.9 (21.0-29.3) |  |
| ≥2 | 48 | 14.2 (10.7-18.4) | 34 | 7.7 (5.4-10.6) |  |
| Specific comorbidities |  |  |  |  |  |
| Immunosuppression | 13 | 3.8 (2.1-6.5) | 22 | 5.0 (3.2-7.5) | .489 |
| Diabetes mellitus | 20 | 5.9 (3.7-9.0) | 22 | 5.0 (3.2-7.5) | .632 |
| Obesity | 30 | 8.9 (6.1-12.4) | 55 | 12.5 (9.5-15.9) | .132 |
| Cardiovascular disease | 56 | 16.6 (12.8-21.0) | 41 | 9.3 (6.8-12.4) | .003 |
| Malignancy | 12 | 3.6 (1.8-6.1) | 12 | 2.7 (1.4-4.7) | .536 |
| Chronic respiratory diseases | 28 | 8.3 (5.6-11.8) | 23 | 5.2 (3.3-7.7) | .107 |
| Kidney failure | 18 | 5.3 (3.2-8.3) | 9 | 2.0 (0.9-3.8) | .017 |
| Liver disease | 16 | 4.7 (2.7-7.6) | 5 | 1.1 (0.4-2.6) | .003 |
| Vaccination status prior to infection |  |  |  |  |  |
| Fully vaccinated<sup>b</sup> | 266 | 78.7 (73.9-82.9) | NA |  |  |
| Partially vaccinated | 23 | 6.8 (4.4-10.0) | NA |  |  |
| Non-vaccinated | 49 | 14.5 (10.9-18.7) | NA |  |  |

Abbreviations: COVID-19, coronavirus disease 2019; NA, not applicable.
<sup>a</sup> 95% CIs were calculated using Clopper-Pearson method.
<sup>b</sup> Individuals were considered fully vaccinated if they had received the required dose(s) of a SARS-CoV-2 vaccine and were at least 14 days after completion.

**Table 2.** Characteristics and prevalent symptoms in patients with mild-to-moderate COVID-19 during the proxy Omicron period versus comparator period.

|  | Proxy Omicron period<br>(n=338) |  | Comparator period<br>(n=441) |  |  |
| --- | --- | --- | --- | --- | --- |
|  | n | Prevalence % (95% CI)<sup>a</sup> | n | Prevalence % (95% CI)<sup>a</sup> | *p-value* |
| Symptoms based on the ARTIQ<sup>b</sup> |  |  |  |  |  |
| Dry cough | 192 | 56.8 (51.3-62.2) | 199 | 45.1 (40.4-49.9) | .002 |
| Coughing up mucus | 88 | 26.0 (21.4-31.1) | 56 | 12.7 (9.7-16.2) | <.001 |
| Blocked nose | 231 | 68.3 (63.1-73.3) | 116 | 26.3 (22.2-30.7) | <.001 |
| Fever | 199 | 58.9 (53.4-64.2) | 295 | 66.9 (62.3-71.2) | .024 |
| Headache | 186 | 55.0 (49.6-60.4) | 200 | 45.4 (40.6-50.1) | .005 |
| Sore throat | 172 | 50.9 (45.4-56.3) | 113 | 25.6 (21.6-30.0) | <.001 |
| Muscle pain | 173 | 51.2 (45.7-56.6) | 223 | 50.6 (45.8-55.3) | .885 |
| Joint pain | 151 | 44.7 (39.3-50.1) | 217 | 49.2 (44.4-54.0) | .219 |
| Chest pain | 72 | 21.3 (17.1-26.1) | 92 | 20.9 (17.2-25.0) | .929 |
| Sinonasal pain | 68 | 20.1 (16.0-24.8) | 54 | 12.2 (9.3-15.7) | .004 |
| Loss of appetite | 96 | 28.4 (23.7-33.5) | 176 | 39.9 (35.3-44.6) | <.001 |
| Problems breathing | 79 | 23.3 (19.0-28.3) | 102 | 23.1 (19.3-27.4) | .932 |
| Wheezing | 48 | 14.2 (10.7-18.4) | 57 | 12.9 (9.9-16.4) | .672 |
| Shortness of breath | 92 | 27.2 (22.5-32.3) | 138 | 31.3 (27.0-35.8) | .235 |
| Felt tired | 241 | 71.3 (66.2-76.1) | 301 | 68.3 (63.7-72.6) | .388 |
| Symptoms (*ad hoc* questions) |  |  |  |  |  |
| Red eyes | 27 | 8.0 (5.3-11.4) | 71 | 16.1 (12.8-19.9) | <.001 |
| Diarrhoea | 70 | 20.7 (16.5-25.4) | 158 | 35.8 (31.3-40.5) | <.001 |
| Nausea | 63 | 18.6 (14.6-23.2) | 80 | 18.1 (14.7-22.1) | .926 |
| Vomit | 15 | 4.4 (2.5-7.2) | 27 | 6.1 (4.1-8.8) | .339 |
| Abdominal pain | 53 | 153.7 (12.0-20.0) | 54 | 12.2 (9.3-15.7) | .174 |
| Insomnia | 62 | 18.3 (14.4-22.9) | 100 | 22.7 (18.8-26.9) | .154 |
| Dizziness | 37 | 10.9 (7.8-14.8) | 55 | 12.5 (9.5-15.9) | .576 |

Abbreviations: COVID-19, coronavirus disease 2019; ARTIQ, Acute Respiratory Tract Infection Questionnaire.
<sup>a</sup>95% CIs were calculated using Clopper-Pearson method.
<sup>b</sup>Prevalence is combined response of “a little” or “a lot”

**Table 3.** Characteristics of the chemosensory dysfunction in patients with mild-to-moderate COVID-19 during the proxy Omicron period versus comparator period.

|  | Proxy Omicron period<br>(n=338) |  | Comparator period<br>(n=441) |  |  |
| --- | --- | --- | --- | --- | --- |
|  | n | Prevalence % (95% CI)<sup>a</sup> | n | Prevalence % (95% CI)<sup>a</sup> | *p-value* |
| Chemosensory impairment (SNOT-22 ≥1) |  |  |  |  |  |
| Yes | 110 | 32.5 (27.6-37.8) | 295 | 66.9 (62.3-71.3) | <.001 |
| No | 228 | 67.5 (62.2-72.4) | 146 | 33.1 (28.7-37.7) |  |
| Type of Chemosensory impairment |  |  |  |  |  |
| Smell | 83 | 24.6 (20.1-29.5) | 276 | 62.6 (57.9-67.1) | <.001 |
| Taste | 91 | 26.9 (22.3-32.0) | 253 | 57.4 (52.6-62.0) | <.001 |
| Smell & taste | 65 | 19.2 (15.2-23.8) | 234 | 53.1 (48.3-57.8) | <.001 |
| Only smell | 18 | 5.3 (3.2-8.3) | 42 | 9.5 (7.0-12.7) | .003 |
| Only taste | 26 | 7.7 (5.1-11.1) | 19 | 4.3 (2.6-6.6) | .371 |
| Severity of alteration of sense of smell or taste (SNOT-22) |  |  |  |  |  |
| 0=None | 228 | 67.5 (62.2-72.4) | 146 | 33.1 (28.7-37.7) | <.001 |
| 1=Very mild | 21 | 6.2 (3.9-9.3) | 7 | 1.6 (0.6-3.2) |  |
| 2=Mild/light | 28 | 8.3 (5.6-11.8) | 35 | 7.9 (5.6-10.9) |  |
| 3=Moderate | 27 | 8.0 (5.3-11.4) | 48 | 10.9 (8.1-14.2) |  |
| 4=Severe | 16 | 4.7 (2.7-7.6) | 64 | 14.5 (11.4-18.2) |  |
| 5=As bad as it can be | 18 | 5.3 (3.2-8.3) | 141 | 32.0 (27.6-36.5) |  |

Abbreviations: COVID-19, coronavirus disease 2019; SNOT-22, Sino-Nasal Outcome test 22.
<sup>a</sup>95% CIs were calculated using Clopper-Pearson method.

### Differences in clinical presentation comparing the two periods

The study cohort was compared with an historical cohort of 441 patients who developed SARS-CoV-2 infection during between March and April, 2020 (comparator period). The two cohorts showed similar distribution by gender, age and smoking status. Approximately one third of patients reported comorbidities in both periods (34.3% in the proxy Omicron period and 32.7% in the comparator period). However, multimorbidity was more frequent in the proxy Omicron period than in the comparator period (14.2% vs 7.7%, *p*=.008). Cardiovascular disease was significantly most frequent in the Omicron period (16.6% vs 9.3%, *p*=.003).

Significant differences in the prevalence of symptoms between the two period were observed (**Table 2**). Particularly, blocked nose (68.3%% vs. 26.3%; *P*<.001), dry cough (56.8% vs. 45.1%; *p*=.002), headache (55.0% vs. 45.4%; *p*=.005), sore throat (50.9% vs. 25.6%; *p*<.001), coughing up mucus (26.0% vs. 12.7%; *p*<.001), and sinonasal pain (20.1% vs. 12.2%; *p*=.004) were more common in the proxy Omicron period, while loss of appetite, diarrhoea, and red eyes were significantly reported more frequently in the comparator period (**Table 2**).

The prevalence of self-reported chemosensory dysfunction during the proxy Omicron period (32.5%) was significantly lower from that during the comparator period (66.9%) (*p*<.001). 24.6% of patients reported an altered sense of smell during the proxy Omicron period compared to 62.6% during the comparator period (*p*<.001). Similarly, the prevalence of an altered sense of taste dropped from 57.4% during the comparator period to 26.9% during the proxy Omicron period (*p*<.001). Moreover, the severity of chemosensory dysfunction, as measured by SNOT-22 score, was significantly lower in proxy Omicron period compared to comparator period (*p*<.001).

### Variables associated with chemosensory dysfunction

None of the tested variables emerged as significantly associated with chemosensory alteration in patients who contracted the infection during the proxy Omicron period (**Supplementary Table**). Vaccination status was not predictive of the chemosensory outcome with 33.3% and 32.3% of fully-vaccinated and partially-vaccinated/unvaccinated subjects, respectively, self-reporting a SNOT-22 ≥1 (*p*=.888). Although nasal obstruction was present in more than two thirds of patients, the prevalence of smell dysfunction in patients with and without nasal obstruction was 25.1% (58/173) and 24.3% (26/81), respectively (*p*=1.000).

## Discussion

We observed a statistically significant reduction in the prevalence of smell and taste alterations in patients who developed the disease during the proxy Omicron period compared to that observed in patients who contracted SARS-CoV-2 infection during the comparator period with the prevalence of smell and taste dysfunction dropping from 63% to 25% and from 57% to 27%, respectively.

One of the possible reasons for this difference is the modulation that the vaccine may have had on clinical expression of SARS-CoV-2 infection. Indeed, vaccination has amply demonstrated its effectiveness in making the clinical manifestations of COVID-19 less severe ^16,17^. However, in the present series the prevalence of chemosensory dysfunction was not influenced by the vaccination status. Furthermore, a vaccination effect on the prevalence of chemosensory disorders does not appear to be supported by several other observations. Current vaccines against SARS-CoV-2 are based on systemic injection which predominantly induces production of circulatory IgG and, potentially, cytotoxic T cells, while are poorly effective at generating mucosal immune responses, i.e. secretory IgA ^18^. Therefore, the olfactory neuroepithelium appears theoretically still vulnerable to SARS-CoV-2 even in vaccinated patients. Early studies found no significant correlation between serum immunoglobulin levels and duration of olfactory disfunction ^19,20^. The correlation is instead significant with nasal immunoglobulin ^20^. Also, vaccination was demonstrated to be less effective against the highly mutated Omicron variant^21^ and the data of the present analysis support this: even patients who received the booster dose developed a symptomatic disease. Finally, we previously observed that chemosensory dysfunctions were among the most frequent symptoms of COVID-19 in vaccinated subjects when the pandemic was mainly driven by the Delta variant ^22^.

The Omicron variant is a highly mutated strain of SARS-CoV-2 showing many substitutions in the spike glycoprotein which may impact on the affinity for the angiotensin converting enzyme 2 (ACE-2) receptor. It has been showed that the supporting cells as well as horizontal basal cells and globose basal of the olfactory neuroepithelium are targeted by SARS-CoV-2 ^23,24^. Both these cell populations display the molecular makeup that makes these cells prone to SARS-CoV-2 infection, i.e., ACE2 receptor and transmembrane serine protease 2 (TMPRSS2), which are, conversely, not expressed by the olfactory sensory neurons ^23,24^. Recent experimental observations support the fact that the Omicron variant has an unique mechanism of cellular entry which shifted cell trophism from TMPRSS2-expressing cells ^25^. Thus, a different interaction between the virus and cellular targets may be responsible for the reduction in the chemosensory impairment observed during the proxy Omicron period.

Smell and taste impairment were consistently described as pathognomonic manifestations of the COVID-19 with several studies confirming the high sensitivity and specificity of self-reported new onset of smell and/or taste impairment for COVID-19 in populations of patients with flu-like symptoms ^26–28^. It is very likely that the advent of Omicron may deprive us of this differential diagnosis tool and that COVID-19 at least in its mild-moderate form can easily confuse with other respiratory infections. Furthermore, symptoms of upper airway involvement, i.e. blocked nose, sore throat, and coughing up mucus, were predominant in patients of the proxy Omicron period and quite more frequent than what observed in patients infected during the first wave of the pandemic.

Regarding the pathogenesis of the alteration of smell, the higher prevalence of nasal obstruction and the less severity of the chemosensory dysfunction observed in the study cohort suggests that at least in part a conductive loss blocking inspired odorants from reaching the olfactory cleft in the nasal cavity could be a possible cause of the loss of smell in patients infected by Omicron variant. However, the overlapping prevalence of chemosensory dysfunction in patients with and without nasal obstruction suggests that, in a fraction of cases, a sensorineural dysfunction of the olfactory neuroepithelium still remains a plausible hypothesis for olfactory impairment, as was previously observed in patients infected by non-Omicron strains ^23^.

The high prevalence of COVID-19 associated chemosensory dysfunction observed in these last two year was unprecedented. The downsizing of the prevalence of these disorders observed with the advent of the Omicron variant should not induce national health services to reduce the resources allocated to the diagnosis and treatment of smell and taste alterations. There are, indeed, a huge number of patients with a long-term COVID-19 dominated by chemosensory alterations^14,29^ with important implications on the quality of life of these subjects ^29,30^. Moreover, even if the prevalence of chemosensory disorders caused by Omicron variant appears reduced, the greater spread of the virus can still lead to a significant number of patients with alterations in smell or taste. It will be of paramount importance to collect data relating to the evolution of Omicron-related chemosensory disorders, i.e. recovery and persistence rate, to fully estimate the burden of chemosensory dysfunction caused by the SARS-CoV-2 Omicron variant.

Finally, the higher prevalence of patients with cardiovascular, hepatic, renal disease, and multimorbidity observed in the study cohort may be due to the lower aggressiveness of the Omicron variant^31^, where patients with these comorbidities tended to develop severe COVID-19 when infected by other SARS-CoV-2 strains^32^.

This study has the following limitations. First, hospitalized patients were not included in the study. Although this made our cohort more homogeneous, studies evaluating the impact of chemosensory dysfunction in more severe Omicron driven COVID-19 are needed. Symptoms were self-reported and based on telephone interview. Although we tried to perform a comprehensive symptoms assessment, some symptoms may have been undetected. Furthermore, a more precise evaluation of the chemosensory function by psychophysical assessment was lacking. Another limitation may be the heterogeneity in the vaccine status across participants. Some of them having one dose of vaccine, while other completed the 3 doses at different times before the conduction of the study. Ultimately, patient inclusion in the proxy omicron period was based on epidemiological data from small samples sequenced regionally. We are therefore unable to estimate to what extent the sample is contaminated by non-Omicron cases. However, to reduce this bias, we decided to limit the analysis to cases of SARS-CoV-2 infection diagnosed after January 17, 2022 when the Omicron variant was estimated to be above 95%.

## Conclusion

The prevalence and the severity of COVID-19 associated smell and taste dysfunction has dropped significantly with the advent of the Omicron variant. Although nasal obstruction was a symptom observed more frequently in the study cohort, the prevalence of chemosensory changes was similar in subjects with and without blocked noses, suggesting that a conductive loss may be the cause of the disturbance only in a fraction of cases. Studying the evolution of chemosensory loss will be of critical importance in assessing the burden of chemosensory dysfunction caused by the SARS-CoV-2 Omicron variant.

## Data Availability

All data produced in the present study are available upon reasonable request to the authors

## Availability of data and material

The authors confirm that the data supporting the findings of this study are available within the article

## Code availability

not applicable

## Author Contributions

Drs Boscolo-Rizzo and Vaira had full access to all of the data in the study and take responsibility for the integrity of the data and the accuracy of the data analysis

*Concept and design:* Drs Boscolo-Rizzo and Vaira

*Acquisition, analysis, or interpretation of data:* All authors

*Drafting of the manuscript:* Drs Boscolo-Rizzo and Vaira

*Critical revision of the manuscript for important intellectual content:* All authors

*Statistical analysis*: Dr Polesel

*Supervision*: Drs Boscolo-Rizzo, Tirelli, Hopkins, Lechien, De Riu, Vaira

## Acknowledgments

We thank Emilia Cancellieri, MD, Andrea D’Alessandro, MD, Rebecca De Colle, MD, Riccardo Marzolino, MD, Anna Mascherin, MD, Chiara Lazzarin, MD, and Enrico Zanelli, MD, for helping in the collection of patient data

## Ethics approval

The study was approved by the Ethics Committees of the Friuli Venezia Giulia Region (CEUR-OS156) and University Hospital of Cagliari (PG 2021/7118)

## Informed consent

informed consent was verbally obtained from all individual participants in the study

---

**Supplementary Table.** Variables associated with chemosensory dysfunction in patients with mild-to-moderate COVID-19 during the proxy Omicron and comparator period.

|  | Proxy Omicron period<br>(n=338) |  |  |  | Comparator period<br>(n=441) |  |
| --- | --- | --- | --- | --- | --- | --- |
|  | SNOT-22≥1 |  | SNOT-22=0 |  | OR (95% CI)<sup>a</sup> | OR (95% CI)<sup>a</sup> |
|  | N | (%) | N | (%) |  |  |
| Sex |  |  |  |  |  |  |
| Male | 45 | (29.0) | 110 | (71.0) | Ref. | Ref. |
| Female | 65 | (35.5) | 118 | (64.5) | 1.27 (0.79-2.02) | 1.54 (1.02-2.34) |
| Age at SARS-Cov-2 positivity (years) |  |  |  |  |  |  |
| <45 | 59 | (37.6) | 98 | (62.4) | Ref. | Ref. |
| 45-54 | 22 | (32.4) | 45 | (67.7) | 0.80 (0.44-1.47) | 0.70 (0.42-1.18) |
| ≥55 | 29 | (25.7) | 84 | (74.3) | 0.60 (0.35-1.02) | 0.62 (0.38-1.00) |
| Smoking status |  |  |  |  |  |  |
| Never | 62 | (31.0) | 138 | (69.0) | Ref. | Ref. |
| Ever | 48 | (34.8) | 90 | (65.2) | 1.27 (0.79-2.04) | 0.97 (0.64-1.48) |
| Comorbidity |  |  |  |  |  |  |
| None | 73 | (32.9) | 149 | (67.1) | Ref. | Ref. |
| 1 | 19 | (27.9) | 49 | (72.1) | 0.97 (0.52-1.82) | 1.56 (0.95-2.57) |
| ≥2 | 18 | (37.5) | 30 | (62.5) | 1.81 (0.87-3.72) | 1.08 (0.51-2.30) |
| Previous SARS-CoV-2 infection |  |  |  |  |  |  |
| No | 106 | (33.0) | 215 | (67.0) | Ref. |  |
| Yes | 4 | (23.5) | 13 | (76.5) | 0.53 (0.17-1.70) |  |
| Vaccination (doses) |  |  |  |  |  |  |
| Unvaccinated/partially vaccinated | 24 | (33.3) | 48 | (66.7) | Ref. |  |
| Fully vaccinated | 86 | (32.3) | 180 | (67.7) | 0.91 (0.52-1.60) |  |
| Blocked nose (smell impairment only) |  |  |  |  |  |  |
| No | 26 | (24.3) | 81 | (75.7) | Ref. | Ref. |
| Yes | 58 | (25.1) | 173 | (74.9) | 0.97 (0.56-1.66) | 1.30 (0.83-2.04) |

Abbreviations: COVID-19, coronavirus disease 2019; SNOT-22, Sino-Nasal Outcome test 22; SARS-CoV-2, severe acute respiratory syndrome coronavirus 2.
<sup>a</sup>95% CIs were calculated using Clopper-Pearson method.

